# Rapid, Reliable and Robust approach for extraction-free RT-PCR based detection of SARS-CoV-2 in clinical setting to expedite large scale screening

**DOI:** 10.1101/2021.11.11.21266209

**Authors:** Abhilasha Dubey, Sanjay Upadhyay, Manjeet Mehta

## Abstract

Rapid, reliable and robust method for the detection of SARS-CoV-2 is an indispensable need for diagnostics. The development of diagnostic method will aid to address further waves of the pandemic potentially with rapid surveillance of disease; and to allay the fears. To meet this challenge, we have developed a rapid RT-qPCR method for the detection of 3 target genes or confirmatory genes in less than 30 minutes. The assay showed 100% sensitivity and 100% specificity when tested on 120 samples. We compared a conventional extraction based method with extraction-free method, and then further reduced the run time of extraction free method. Additionally, we have validated our rapid RT-qPCR method for the assessment of pooled sample. We hereby propose a most reliable approach for the mass screening of samples with ease of operation at low cost. Finally we designed a single tube analysis method which provides qualitative as well as quantitative results in minimum time.

## Background

The emergence of two consecutive waves have given rise to a concern for the world’s 2^nd^ largest populated country to establish a rapid, relay and reliable method for SARS-CoV 2 detection for mass screening. The current need of the hour is to develop large scale diagnostic method to determine the spread of the virus in Indian population quickly compressively, precisely, with high security & specificity. The widely used golden most reliable approach for the detection of SARS-CoV 2 in clinical diagnostic is an RT PCR assay which is based on Nucleic Acid Amplification Testing NAAT. The new approaches under NAAT are emerging for rapid detection of disease. CDC has approved NAAT based methods to be used for point of care diagnosis for mass screening. Recent attempts have been made to circumvent RNA extraction in SARS-CoV-2 detection. Herewith we established new approach for SARS-CoV 2 RNA detection by extraction free single tube detection RT PCR for SARS-CoV 2.

## Introduction

Increase in Occurrence of pneumonia like cases has created a scourge circumstance in nationwide China in December 2019. In lap of time within a period of month, this epidemic changed into a terrifying pandemic and started spreading to different nations via movement of people [1]. In January 2020, World Health Organization (WHO) showed concern about arising endemic situation and issued regulatory guidelines. In March WHO declared 2019-CoV as a pandemic and affirmed an international health emergency. WHO reported that this pandemic is because of novel coronavirus (2019-CoV) that causes disease known as COVID-19 [2]. On 11^th^ February 2020 International Committee on Taxonomy of Viruses named 2019-nCoV as severe acute respiratory syndrome coronavirus 2 (SARS-CoV-2) [3]. As per WHO info reported in September 2021, 237,221,311 confirmed cases, 214,364,453 recoveries, and 4,843,447 deaths were reported in around 223 countries areas and territories [4]. India being the second most populated country after China faces a great threat due to novel CoV and reported 33,894,312 confirmed cases, 33,200,258 recoveries, and 449,883 deaths were reported in around [5].

The genome of the novel coronavirus SARS-CoV-2, causing atypical pneumonia in human population of Wuhan, had 89% nucleotide identity with bat SARS-like CoV ZXC21 and 82% with that of human SARS-CoV-2 and proved this as a new virus strain called SARS-CoV-2 or COVID-19 [6]. Phylogenetic tree analysis using the whole genome sequences of SARS-CoV-2 with five severe SARS-CoV-2 sequences, two MERS-CoV sequences and five from bat SARS-like coronavirus) from China origin showed that SARS-CoV-2 have formed different cluster and were more similar to Bat SARS-like coronavirus (almost 80%) [7] [8]. Human angiotensin converting enzyme receptor (ACE2 cell receptor) was recognized by both SARS-CoV and Novel coronaviruses SARS-CoV-2 by previously standardized animal models experiments [9]. SARS-CoV-2 genome is made up of spherical or pleomorphic (having diameter of approximately 60–140 nm), single-stranded enveloped RNA molecule covered with club shaped glycoprotein [10] [11]. The genome consists of 29891 nucleotides which encodes 9860 amino acids. The Structural proteins are encoded by the four structural genes, including spike (S), envelope (E), membrane (M) and nucleocapsid (N) genes [12]. Till date there are four sub types are reported of Corona viruses such as alpha, beta, gamma and delta virus. Some of them were affect human of other affected animals such as pigs, birds, cats, mice and dogs [11]. SARS-CoV-2 or COVID-19 belongs to the beta CoVs category. The SARS-CoV-2 can be destroyed by ultraviolet rays and heat. Other lipid solvents including ether (75%), ethanol, chlorine-containing disinfectant, peroxyacetic acid and chloroform can also effectively inactivate this virus particle [13].

SARS-CoV-2 belongs to the coronavirus genus β with a single stranded, non-segmented positive-sense RNA genome, which is the seventh known coronavirus that can infect humans [14][15]. As related to other morbific RNA viruses, the genetic RNA material is the most reliable diagnostic marker to be detected. The nucleic acid amplification test (NAAT) is currently used in conjunction with pulmonary Computed Tomography (CT) for the clinical diagnosis of COVID-19. Sequencing and mutational analysis are carried to find out hot spot mutation for the surveillance of new variants [16]. After onset of infection, antibodies IgM and IgG are produced by the human immune system in response to antigens with prognosis of disease. Lateral flow based test based on antigen antibody reaction plays a major role in monitoring the response the effect of immunization and also reveals previous exposure/immunity, the antigen antibody based detection always lag behind sensitivity and specificity of conventional RTqPCR [18].

The current RT-PCR nucleic acid detection are also subjected to false positive and negative results in some cases. The confirmatory or target genes for SARS-CoV-2 are based on the conserved and specific sequences of viral genome like open reading frame 1ab (ORF1ab), spike (S),RNA-dependent RNA polymerase (RdRp), envelope (E), and nucleocapsid (N) genes [14]. Although ORF1ab is the highest specificity confirmation target gene, it is considered to be less sensitive than other targets in clinical application. For large scale screening of disease pursuance of mixed sample testing or pooling was approved by CDC and Government of India https://www.mohfw.gov.in/pdf/GuidelineforrtPCRbasedpooledsamplingFinal.pdf, https://www.cdc.gov/coronavirus/2019-ncov/lab/pooling-procedures.html. However, some reports propose that although this method improves the detection efficiency, it might increase incidence of false negative results or individuals with low viral load [19] [20]. In addition, currently approved nucleic acid test kits do not mention whether they could be used for mixed sample detection. Thus clinically,it is recommended that samples with suspicious results or single channel positive results should be re-examined with another manufacturer’s kit or method. Nonetheless, the basis for selecting a specific validation kit is still under development.

As per reference data available by World Health Organization (WHO), the SARS-CoV-2 can stay in infectious stage for up to 72 hours on plastic and stainless steel, less than 4 hours on copper, and less than 24 hours on cardboard [21]. However, the virus can be detected in environments after this time, indicating that environmental conditions like temperature, for example, can inactivate the viral particles but, in the meantime, is not able to entirely degrade viral genetic material, making it possible to detect it. SARS-CoV-2 can be heat inactivated at 60°C, 80°C, and 100°C for approximately 32.5, 3.7, and 0.5 minutes, respectively, and these times and temperatures held enough to reduce the infectivity of virus[22]. The same range of temperatures are also used in an amplification program of RT-PCR, indicating that the genetic material’s quality is not compromised. Furthermore, using an internal control for Real time RT-PCR reaction can confirm the samples’ integrity, decreasing the probability of false-negative results, Based on this knowledge, physical methods like heating and specific chemical reagents like proteinase K and other RNase inhibitors are widely incorporated in extraction protocols to optimize viral purification.

To meet current demands in diagnostic setting for rapid detection of disease, we established a protocol using one step RT-PCR test from nasopharyngeal samples in Universal Transport Medium (UTM) followed by thermal shock for a faster and low-cost RNA extraction for COVID-19 diagnosis. We compared our new method’s effectiveness based on (cycle threshold) Ct values obtained by Real time RT-PCR specific for SARS-CoV-2 using a constitutive human gene (RNaseP) as an internal control to insure human specimen collection for the test.

## Materials and methodology

### Ethics statement

All methods were carried out in accordance with relevant guidelines and regulations. Assays were performed on existing anonymised RNA samples collected during standard diagnostic tests at our laboratory registered with National Accreditation Board for Testing and Calibration Laboratories (NABL) registration no. MC-4413 & approved by Indian council of medical research (ICMR) for SARS-CoV-2 testing (ICMR-MEDCMMH), with no clinical or epidemiological data available, apart from each reported quantification cycle (Cq) or cycle threshold (Ct). Consent was not required as stored samples that have been taken for diagnosis and remain after the diagnostic procedure, providing that all samples are anonymised to researcher.

### Sample collection

Clinical samples (nasopharyngeal and oropharnygeal) swabs were collected in a tube containing viral transport medium on site and at Medilab diagnostic center, molecular biology laboratory by the expert phlebotomist. A total of 150 known positive and negative samples were reassessed for developing and validating an extraction free feasible method. For RNA purification for extraction based method, 200μl of sample from virus transport medium was extracted using HiPurA™ Viral RNA Purification Kit (#MB605) followed by elution and purification using Magnetic Bead based Automated Nucleic acid Extractor (Hi-media Insta NX Mag 32). In this study we established extraction free method for detection of SARS-CoV-2 by using direct lysis buffer. The differences in Cycle threshold (Ct) was compared between both extraction based and extraction free RT-PCR for validating the results. The direct detection method used in this study is simple, economical, and robust method was developed for mass screening of samples.

### RNA Purification from clinical samples

For RNA extraction, 200μl of samples was purified using HiPurA™ Viral RNA Purification Kit (#MB605) according to the manufacturer’s protocol. The purified RNA was eluted in provided elution buffer according to Center for Disease Control & Prevention CDC recommendations.

### Heat inactivation

Clinical samples from Nasopharyngeal and oropharnygeal swab in viral transport medium, 100 μl of sample were aliquoted in RNAase free PCR tube was vortexed followed by heat inactivation at 70° C for 5 minutes followed by cooling at 4° C.

### Direct sample with heat inactivation

To skip extraction process of clinical samples, heat inactivated samples were incubated with lysis buffer for 2-3 minutes at room temperature and further mixed with pipetting and sort vortex and used in RT-PCR

### qRT-PCR

We performed qRT-PCR using both extracted RNA and direct lysis of samples, targeting the N1 & N2 gene in the conserved region of SARS-CoV-2 genome. For sampling control for validating the presence of biological samples in SARS-CoV-2 negative samples or positive samples RNAse P was used. A single strand IVT (invitro transcription) RNA was used as an internal control, which helps in verifying presence of contaminants that could inhibit reverse transcription. Final reaction of 25 μl was prepared by mixing 6.25 μl of primer mix, 8.75 μl of master mix and 10 μl of heat inactivated-lysed samples. The thermal cycling steps for 10 minute reverse transcription at 50° C, 2 min of initial denaturation at 95 ° C and 40 cycles of 5 sec denaturation and 30s of annealing & extension at 95 and 60 ° C by using Qiagen artus RT-PCR kit (artus SARS-CoV-2 Prep&Amp UM Kit #4511440). For Meril PCR-kit (Meril COVID-19 One-step RT-PCR Kit #NCVPCR-02), thermal cycling step including 15 minutes of reverse transcription at 50 ° C, initial denaturation for 3 minutes 95 ° C, 40 cycles of 15 sec of denaturation and 40 sec of annealing & extension at 95 & 55 ° C.

Minimum qRT-PCR run times were established preparing a single reaction mix, sufficient to carry out all experiments. The reaction mix consisted of master mix and primers probes specific to target and kept on Ice until required time. 10 μl of samples were added in the same tube and were subjected to qRT-PCR for decreasing. RT-qPCR reactions carried out on the following instruments, Hi-media insta Q96, and Qiagen quant 5 plex thermo cycler. Data were analyzed using instrument software, graph pad prism 5.0, Microsoft excel.

### Sample pooling

On the basis of published article for sample pooling and ICMR guidelines https://www.icmr.gov.in/pdf/covid/strategy/Advisory_on_feasibility_of_sample_pooling.pdf we opted for nasopharyngeal swab pools consisting of 5 combined samples in each well. 10μl of neat sample were taken in PCR tubes followed by heat inactivation at 70° C for 5 minutes, out of which 10 μl was used for final reaction volume. For the positive sample deconvolution technique was used to find out positive samples among the pooled samples.

### Statistical analysis

All data are presented as the mean ± standard error of mean of at least three experimental repeats. Mean values data showed a Gaussian distribution. Comparisons between two groups were performed using a t test. All the statistics were analyzed Graph Pad Prism 5 (Graph Pad Software, Inc.). P<0.05 was considered to indicate a statistically significant difference.

## Results

### Comparison of results using conventional extraction based vs. extraction free method for detection of SARS-CoV-2 using RT-PCR method

A total of 23 samples were reanalyzed for this assay, including 12 positive and 11 negative samples with the standard methodology using the extraction step recommended by (Hi PurA viral purification kit. The mean of quantification cycles (Cq) or cycle threshold (Ct) with total positive (TP) was 26.83 and 26.22 for the N & RNAse P gene, were as N gene ranges from (15.62-33.23) and RNAse P gene ranges from (22.55 – 30.60), respectively shown in table 1.

**Table 1:** Results obtained with the extraction free methods in comparison with conventional approach.

| Kit/ Assay | Method | Positive | Negative | Total | Sensitivity | Specificity |
| --- | --- | --- | --- | --- | --- | --- |
| Meril | Extraction (Ex) | 12 | 11 | 23 | 95% | 95% |
| Qiagen | Ex- free | 11 | 11 | 23 | 92% | 100% |

On the other side in comparison to extraction free method based on heat inactivation, out of total positive samples 12/13 (92%) were positive, respectively, while no total negative samples was called positive using extraction free method.

### Development rapid qRT-PCR for a brisk in sample processing time

In order to develop a potential throughput of the assay in a diagnostic setting, the ability of the assay to perform adequately under short amplification program for both Reverse transcription (RT) and PCR condition was modified to achieve rapid processing rate. In previous experiment we showed that there was no significant change in Cqs value when compared to conventionally used extraction based method to extraction free method, hence we followed extraction free method for sample processing further.

The baseline Cqs data from the standard program setting as per manufacturer (Qiagen artus UM Amp kit) with initial amplification program condition of 10 min RT, 2 min initial denaturation (ID) with PCR setting for 40 cycles of 5 sec denaturation and 30 sec of polymerization were obtained for all three viral targets, showed in figure 2 as standard program. Then subsequently RT time was reduced to 5 min, followed by ID of 1 min and denaturation and polymerization time was bring down to 1 sec and 8 sec for 40 cycles, although including instrument configuration for fluorescence screening required time of 4-5 sec. There was no change in the performance of the assay but the run time was reduced to 37 min 20 sec from 57 min on Qiagen aquant 5plex multichannel detector. This amplification setting conditions were applied to replicate the results.

**Figure 1:**
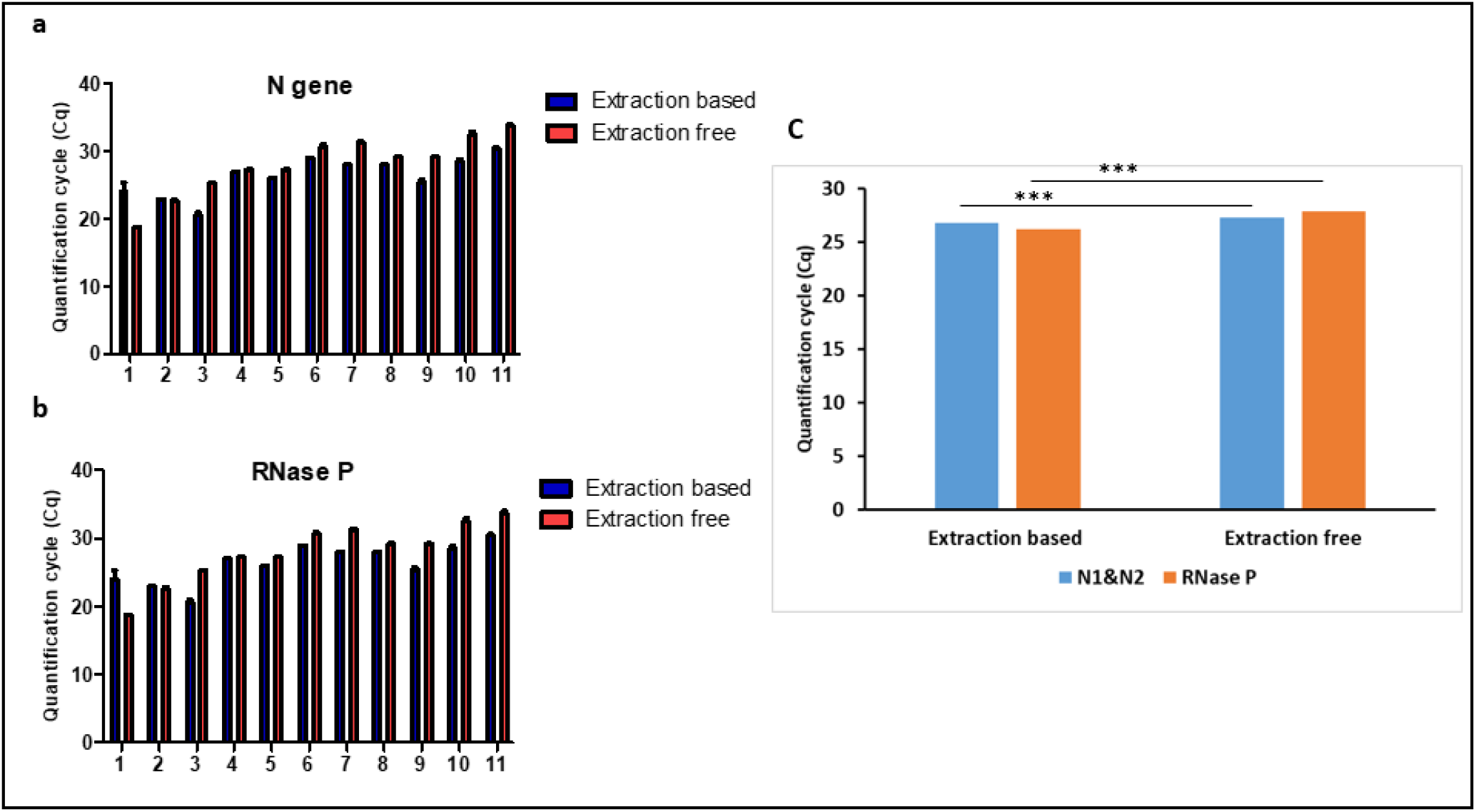
Comparison of Cqs obtained from extraction free method with conventional method.a) Individual Cqs of 11 replicates of patient compared with extraction free method for expression of N gene. b) Comparative Cqs for RNAse p gene for 11 replicate samples of patient.c) Comparative Cqs between extraction based and Ex-free method for confirmatory gens. *** = p<0.0001 significance was calculated by unpaired two-tailed student’s t test.

**Figure 2:**
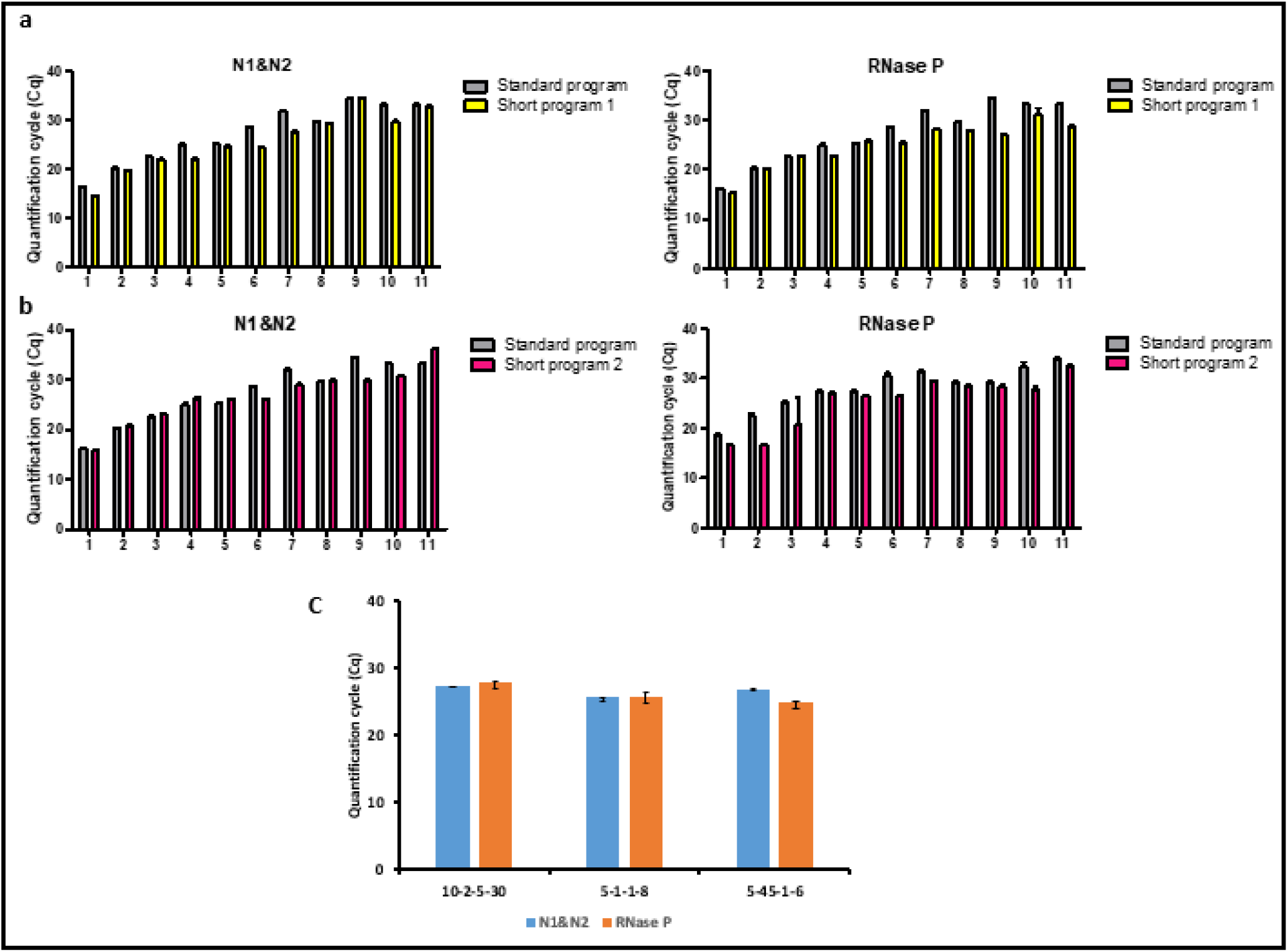
Reduction in qPCR times. a) Cqs obtained for target gens by reduction of qPCR times by reducing RT time and PCR for denaturation to 1 sec and annealing to 8 sec. b) Cqs obtained upon further reduction in qPCR run time by reducing annealing to 2sec and keeping RT at 5 min followed by ID of 45 sec. c) Comparison of initial of RT 5 min, ID 2 min and qPCR of 40 cycles of denaturation 3 sec and annealing 30sec with short program 1 & 2. The assay were run in duplicate, with the plots showing delta Cqs between standard and shorter timings.

The next aim was to reduce run times by further reducing the denaturation and polymerization time. Here the reduction in ID time to 45 sec and annealing time to 6 sec for 40 cycles, while keeping RT time as per previous setting. Upon reducing run time to 33 min 15 sec, there was no decline in assay performance noted. For the reproducibility of this assay we replicated the experiment with more no. of samples. The Cqs from 3 target genes were compared, there was a very little difference with ± 1 deviation with initial result, showed in figure 2.

As the performance of 2 short program of 33 min run time was concordant with repeated validity run, we further aimed to reduce run time further. The cooling step in the block based qPCR instrument is the slowest part, we focused on reducing the temperature gap between denaturation and annealing steps. We further reduced RT to 3 min and ID to 30 sec followed by 40 cycles of denaturation & annealing for 1 sec, we also increased annealing temperature to 62 °c for reducing the cooling time. Upon modification in PCR condition the run time was further reduced to 25 min from 33 min. The sensitivity and specificity of this assay was 95% and 99% respectively, when compared to previously validated assay, shown in figure 3. We repeated experiment with same assay condition for more no. samples to validate this method. Even without any modification to primer or enzyme concentrations, the small change in amplification program, indicated that this method can be potentially used to process sample in short time, as minimum as 25 min.

**Figure 3:**
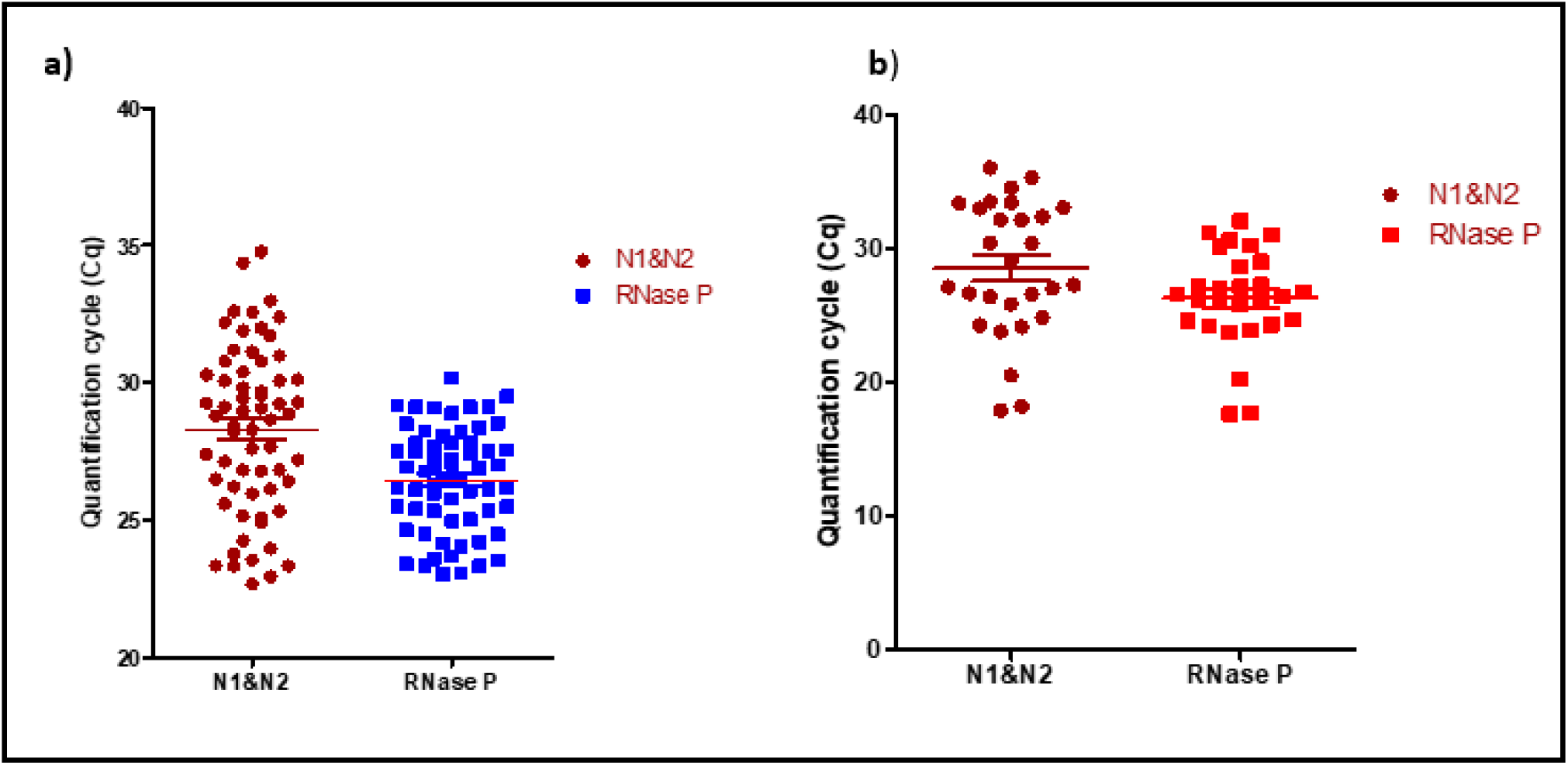
Development of rapid qPCR method. a) Cqs obtained after reducing the temperature gap between denaturation and annealing, with modification in PCR condition, whereas RT was kept for 2 min followed by 1 sec of denaturation and 2 sec of annealing time, which reduced run time to 25 minutes. b) Relative Cqs for previously validated program of 33 min.

**Figure 4:**
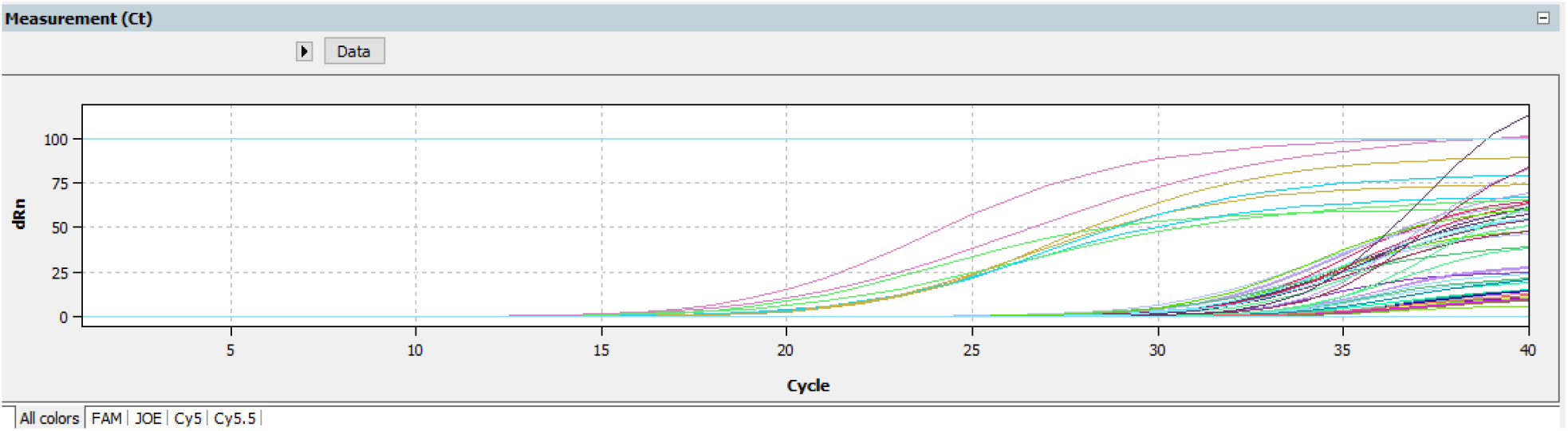
Graphical illustration of pooled sample on rapid PCR setting

**Figure 5:**
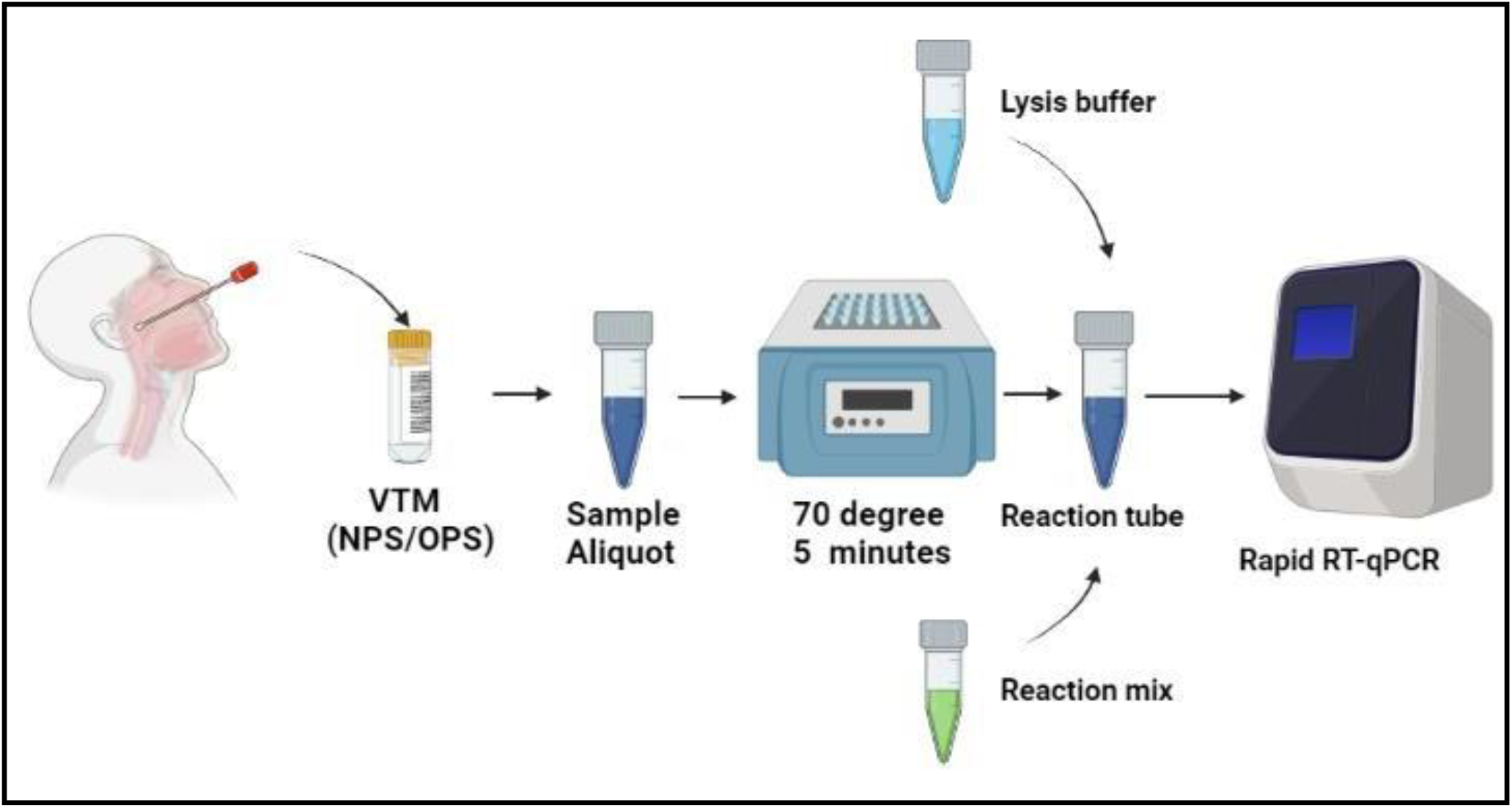
Schematic workflow for single tube reaction format for rapid detection for SARS-CoV-2 detection.

### Analytical sensitivity of rapid qRT-PCR for pooled samples

In order to develop a fast reliable method for mass screening of sample by NAAT using qPCR method, we aimed to analyze sensitivity of our rapid PCR protocol on pooled sample assessment. Here we pooled a total of 100 samples, with 5 samples in each well. Positive signal was detected in 4 wells, which upon deconvolution resulted in a total of 12 positive samples in a batch of 100 specimens. The fast qPCR protocol was used with run time of 25 min. The sensitivity was found to be 99.99 % for this method.

## Discussion

The pandemic, caused by the SARS-CoV-2 virus, has led to the development of a wide range of diagnostic assays, many of which are RT-PCR based which is a confirmatory or golden standard method which, utilize real time detection and report a Cq or ct value to indicate presence or absence of the virus. It has become increasingly clear that there are significant shortcomings in the use and Interpretation of many of these assays for testing and monitoring populations for viral spread. This has resulted in some confusion as to whether these diagnostic assays are capable of adequately addressing their three main functions: First, to identify patients presenting with symptoms consistent with COVID19 as SARS-CoV-2 positive or negative. Second, to provide a meaningful assessment of viral load, given that Cqs are subjective and not sufficiently reproducible or robust to allow an appraisal of the validity of marginal results, i.e. those around cycle 35. Third, to monitor the spread of virus using screening programs of populations and environmental samples, allowing that a high percentage of those infected remain asymptomatic [23].

The current need of hour after facing two subsequent wave of pandemic is to design a diagnostic assay featuring high specificity and sensitivity, with reliability, speed and ease of processing with minimal risk of contamination. In addition, for screening large scale of population cost saving approach is a serious consideration.

In this study we developed a simple, reliable and rapid qRT-PCR method which offers maximum sensitivity by targeting three viral targets with extraction free method. Our aim was to develop a single tube reaction format, with low consumables requirement and less risk of contamination to reproduce test results in less than 30 minutes. The current development in field of NAAT based diagnostic kits for rapid qualitative analysis of disease are limited to process large no. of samples in short time and also the consumables for CB-NAAT or TRU-NAAT are very costly as compared to conventional qPCR tests. Herewith the motivation to develop a diagnostic method for detection of SARS-CoV-2 rapidly as compared other NAAT applications.

Our rapid qPCR method furnished as an adaptable assay that can be applied to all SARS-CoV-2 detection and quantification applications using kits mentioned in this assay. We validated rapid protocol for the processing of pooled sample with run time of 25 mins, which will be very helpful in keeping track of spread for large populations in short time.

In conclusion, with reference to available literature we designed, optimized and validated our rapid quantification and qualitative method as a value-added RT-qPCR assay for SARS-CoV-2. It is robust, rapid and reliable protocol for providing the opportunity for high throughput, multiplex viral detection with the potential to quantify viral load. It is designed to reduce the assay failure and risk of contamination and provides a promise for an alternative assay for point of care testing.

## Data Availability

All data produced in the present work are contained in the manuscript.

## Author’s contributions

A.D. initiated this work and designed all assays, carried out the laboratory work, analyzed the data and drafted this manuscript. MM was the clinical lead and supervised sample selection and helped in drafting manuscript. S.U. assisted the assay design and overviewed the manuscript.

## Consent to participate

Not applicable, Consent was not required as stored samples that have been taken for diagnosis and remain after the diagnostic procedure

